# A structured open dataset of government interventions in response to COVID-19

**DOI:** 10.1101/2020.05.04.20090498

**Authors:** Amelie Desvars-Larrive, Elma Dervic, Nils Haug, Thomas Niederkrotenthaler, Jiaying Chen, Anna Di Natale, Jana Lasser, Diana S. Gliga, Alexandra Roux, Abhijit Chakraborty, Alexandr Ten, Alija Dervic, Andrea Pacheco, David Cserjan, Diana Lederhilger, Dorontinë Berishaj, Erwin Flores Tames, Huda Takriti, Jan Korbel, Jenny Reddish, Johannes Stangl, Lamija Hadziavdic, Laura Stoeger, Leana Gooriah, Lukas Geyrhofer, Marcia R. Ferreira, Rainer Vierlinger, Samantha Holder, Samuel Álvarez, Simon Haberfellner, Verena Ahne, Viktoria Reisch, Vito D.P. Servedio, Xiao Chen, Xochilt María Pocasangre-Orellana, David Garcia, Stefan Thurner

**Author notes:** Corresponding author(s): Amélie Desvars-Larrive.

## Abstract

In response to the COVID-19 pandemic, governments have implemented a wide range of nonpharmaceutical interventions (NPIs). Monitoring and documenting government strategies during the COVID-19 crisis is crucial to understand the progression of the epidemic. Following a content analysis strategy of existing public information sources, we developed a specific hierarchical coding scheme for NPIs. We generated a comprehensive structured dataset of government interventions and their respective timelines of implementation. To improve transparency and motivate collaborative validation process, information sources are shared via an open library. We also provide codes that enable users to visualise the dataset. Standardization and structure of the dataset facilitate inter-country comparison and the assessment of the impacts of different NPI categories on the epidemic parameters, population health indicators, the economy, and human rights, among others. This dataset provides an in-depth insight of the government strategies and can be a valuable tool for developing relevant preparedness plans for pandemic. We intend to further develop and update this dataset on a weekly basis until the end of December 2020.

## Background & Summary

Non-pharmaceutical interventions (NPIs), also known as community mitigation strategies, aim to prevent the introduction of infectious diseases (preparedness and readiness measures), control their spread and reduce their burden on the health system (control measures). The general concept of containing the initial (exponential) spread of a disease is called “flattening the (epi-) curve” ^1^. By reducing the growth rate of an epidemic, NPIs reduce the stress on the healthcare system and help gaining time to develop and produce vaccines and specific medications, which is of utmost importance in the case of emerging infectious diseases.

During the COVID-19 pandemic, governments have enforced a broad spectrum of intervention measures, under rapidly changing, unprecedented circumstances. Government responses to COVID-19 included the *laissez-faire* strategy, which implies doing little to nothing, the *herd immunity* strategy, which implies a few measures only or measures relying on voluntary compliance, and more *aggressive* approaches based on the implementation of a wide range of stringent NPIs, sometimes even limiting civil rights and liberty ^2,3^. Governments control policies have shown divergences in particular in the timeline of implementation and in the prioritization of the NPIs. In China for example, quarantine, social distancing, cordon sanitaire, and isolation of cases have been associated with improvements in the key epidemiological markers, including the number of infections and COVID-19-related deaths ^4^. In Hong Kong and Taiwan, which experienced severe acute respiratory syndrome (SARS) epidemics in 2002-2003 ^5^’^6^, early government actions, strict social distancing measures, contact tracing, extensive and proactive testing, and high compliance of the population, have, to date, successfully mitigated the COVID-19 epidemic ^7,8^. Following a *herd immunity* approach, similar to the one initially adopted by the UK government, the Swedish government did not introduce strict bans but formulated non-binding recommendations only ^9^. Predictive models, however, suggest that such a strategy might ultimately overwhelm the healthcare system due to shortages in critical care capacity ^10^.

Poor control policies have potentially dramatic repercussions on public health. Although there is an urgent need for data on government responses to COVID-19, there is currently a limited opportunity to capture information on country-based mitigation measures. Our project aims to generate a comprehensive and structured dataset of government interventions taken to control the spread of COVID-19, including the respective time schedules of their implementation, which can enable to assess the effectiveness of these interventions and facilitate an inter-country comparison.

During the COVID-19 crisis, other data collection initiatives have emerged on this topic; all of them offering public access to data. Nonetheless, most of them focus on a specific type of measures, e.g. the closure of educational institutions ^11,12^, travel restrictions^13^, trade-related measures^14^, legal and economic responses^15^, or measures to ensure continuity of supply of personal protective equipment and critical medical products^16,17^. Two datasets describe a larger range of interventions; however, the number of measures per country is limited ^18^, measures are not explicitly detailed, or sources are not provided^19^.

In the context of the current COVID-19 health crisis, open knowledge ^20^ and data sharing are crucial to understand and help to mitigate the pandemic. In this article, we document and share the methodologies, tools and approaches used to produce the *Complexity Science Hub COVID-19 Control Strategies List* (CCCSL) dataset following the principles of open science. We provide a detailed description of the dataset and present examples of how the data can be used in order to provide insights into the global government responses to COVID-19. The dataset is readily usable for modelling and machine learning analyses.

By compiling the information on NPIs, the CCCSL dataset allows evaluating the government policies against COVID-19 and should further enable to estimate the impact of these measures on population health indicators, the economy, and human rights, among others. The CCCSL can also be used as a tool for designing emergency preparedness and response plans for epidemic respiratory diseases. Considering the imperative necessity for such data, version 1 of the dataset has already been released. The dataset is not complete and we continuously update it with new available records. Updates are planned until, at least, the end of December 2020.

## Methods

We used a content analysis^21-23^ strategy of existing information sources to develop a hierarchical coding scheme specific to NPIs implemented to mitigate the burden of COVID-19. First, based on a literature review on community mitigation strategies and expert knowledge, eight themes (thereafter called level 1 (L1) in the coding scheme) were identified and labelled: (i) Case identification, contact tracing and related measures, (ii) Environmental measures, (iii) Healthcare and public health capacity, (iv) Resource allocation, (v) Risk communication, (vi) Social distancing, (vii) Travel restriction, and (viii) Returning to normal life. A definition for each theme is provided in Table 1. At the start of our project, there were no previously published studies on NPIs against COVID-19 to be used as a reference for developing our labelling and coding scheme. Therefore, a list of NPIs that have been already implemented by different governments at this time (mid-March 2020) was compiled, that served as a preliminary template to generate *a priori* categories within a hierarchical coding scheme. Strategies that could provide assistance to the population (e.g., related to financial support or food supply) or that may encourage compliance with the measures (e.g. resource allocations, risk communication) were also included. Listed intervention measures were then assigned to one of the eight themes defined above. The specific details and descriptions of each NPI were coded into *a priori* categories (thereafter called level 2 (L2) in the coding scheme), and into subsequent *a priori* subcategories and codes whenever needed (thereafter called level 3 (L3) and level 4 (L4) in the coding scheme, respectively). Discrepancies in code assignments were discussed within the coding team and were resolved by consensus. The objective of this hierarchical coding scheme for NPIs was to standardize the data collection and obtain a structured dataset that uses a consistent taxonomy, and therefore, promotes common understanding.

**Table 1.**
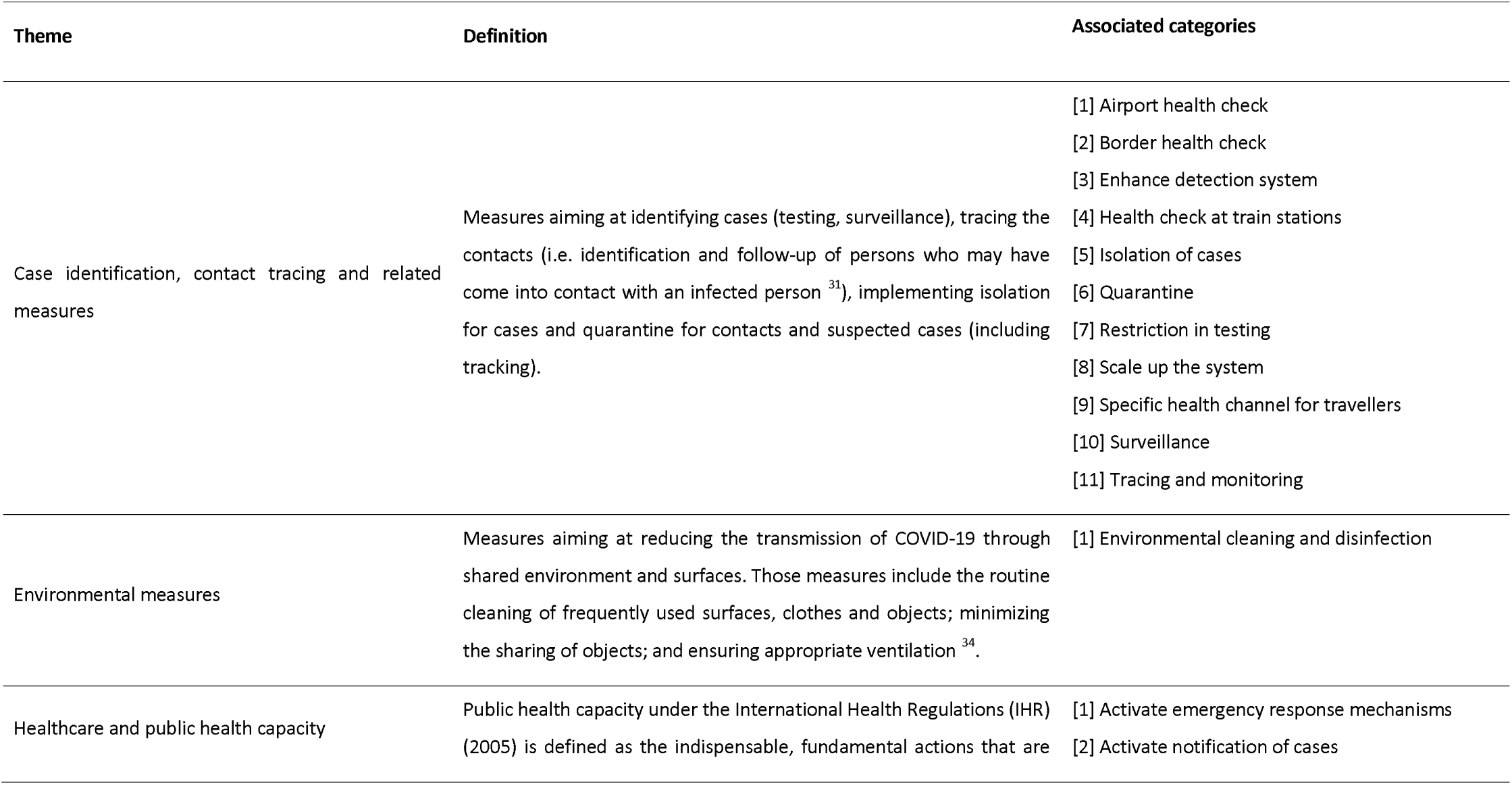
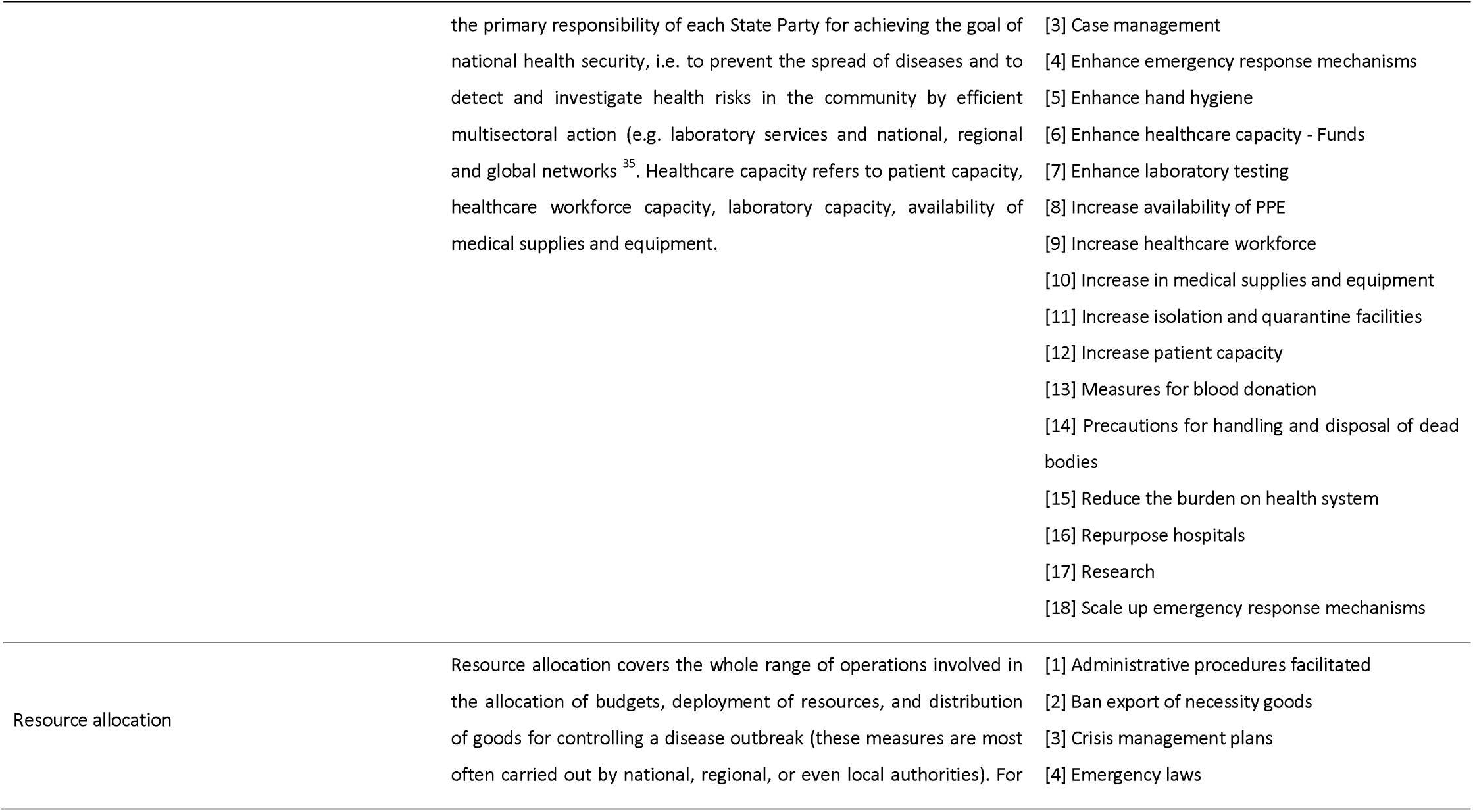
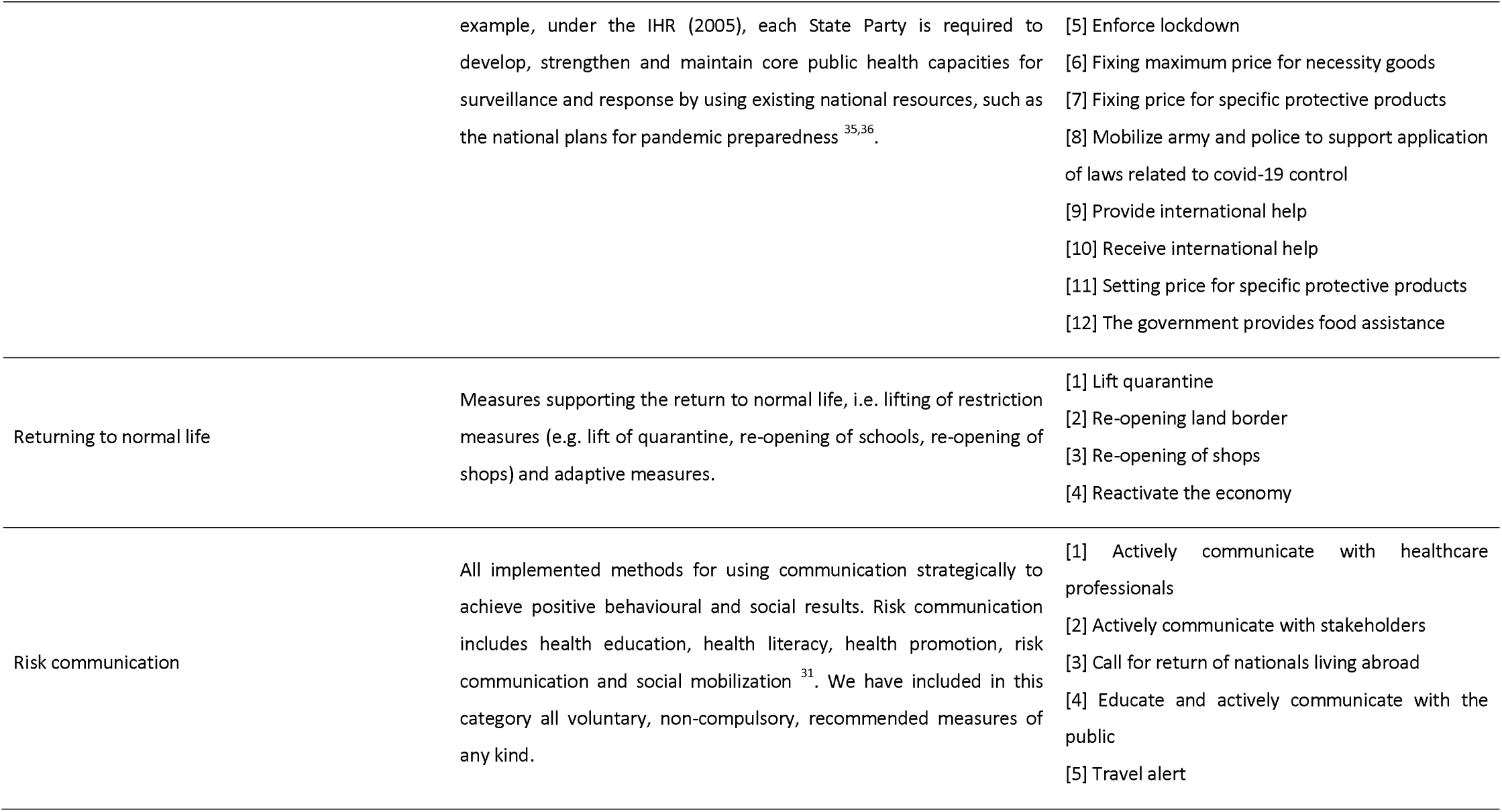
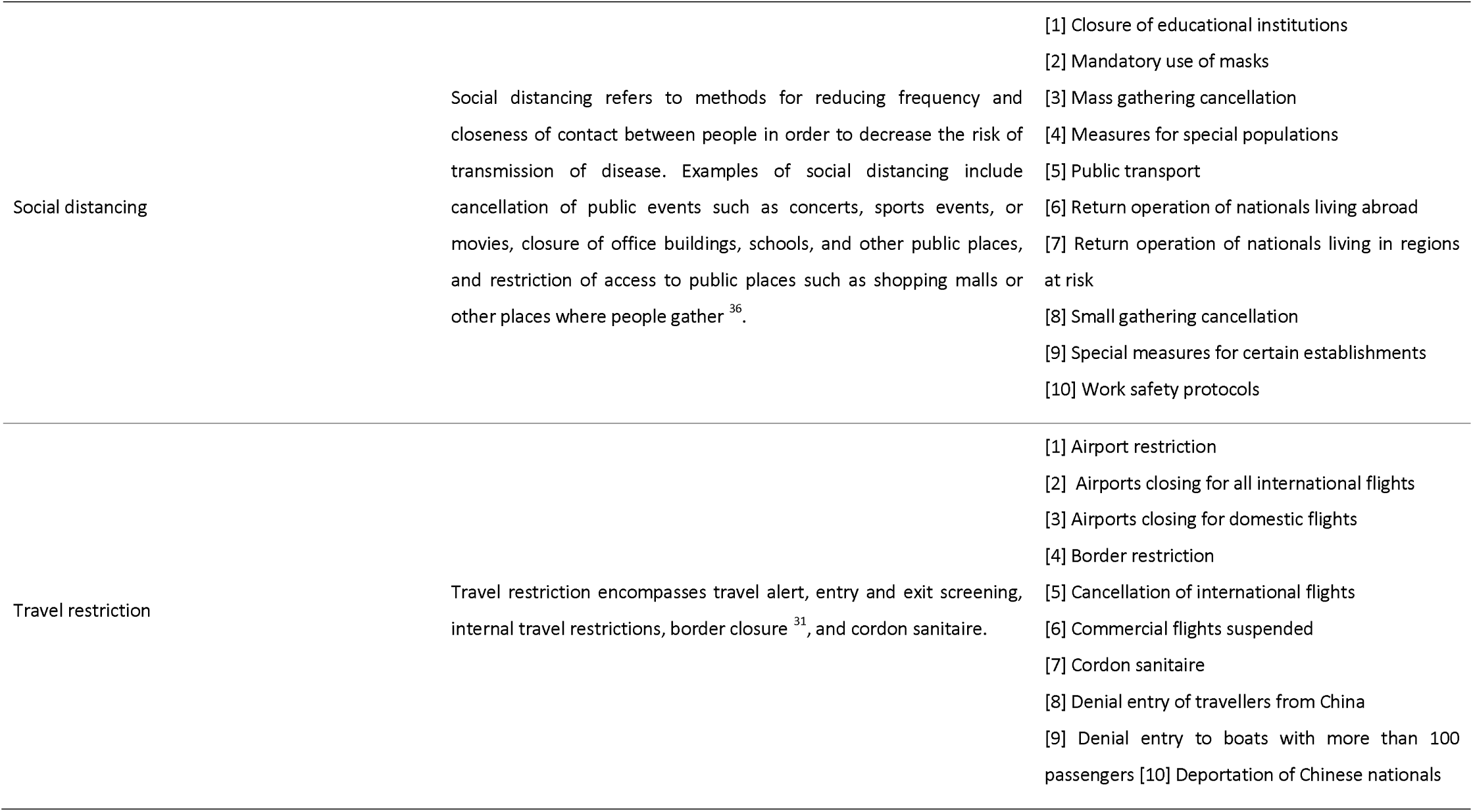
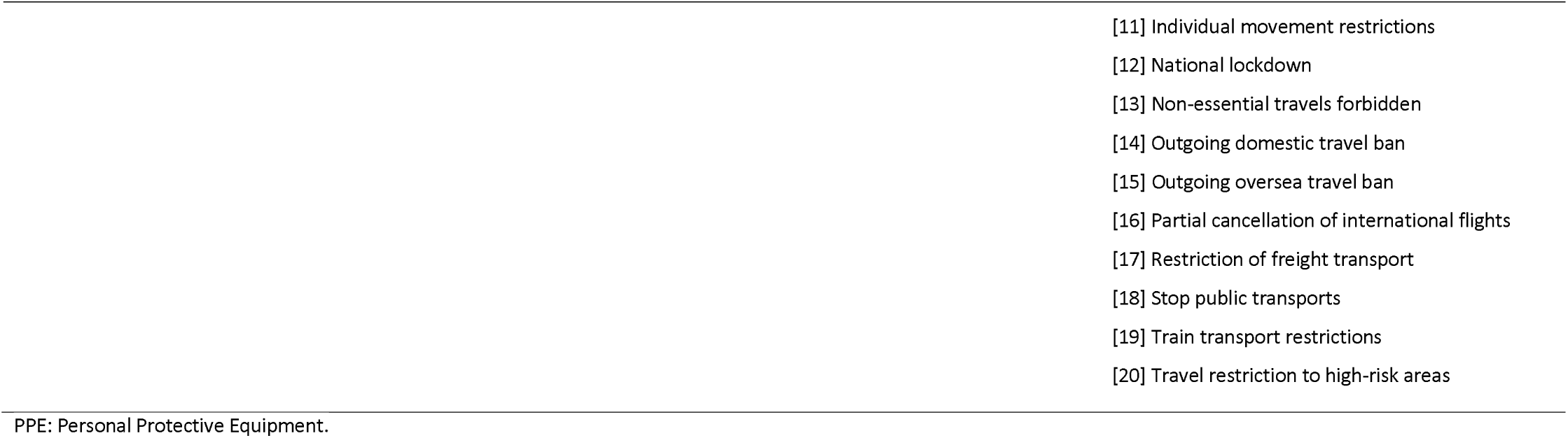
Definition of the eight themes (L1) used to classify the NPIs and associated categories (L2).

On 19 March 2020, we set up a platform for students, researchers, and volunteers to collect data on the NPIs implemented by the governments for preventing and limiting the spread of COVID-19, including the time schedules for the implementation. Data collectors received clear instructions on the objective of the project and indications on how to proceed for data collection. The template of *a priori* themes, categories, subcategories, and codes described above was provided. Data collectors were asked to use the *a priori* coding system or to refer to the data curators if a measure could not be coded using the *a priori* coding system. Therefore, throughout the data collection process, new categories, subcategories, and codes emerged, derived directly from the text data sources. The emergent (inductive) categories and subcategories were openly coded by the data collectors or by the data curators. In a second step, inductive categories and subcategories were compared together and in relation to the entire dataset to detect co-occurrence (codes that partially or completely overlap) and redundancies. Codes with the same meaning were aggregated^24^. The categories and subcategories were tightened up to the point that maximized mutual exclusivity and exhaustiveness^23^. This resulted in a master list of coding categories (a list of all the codes that were developed and used in the study), including the curated *a priori* and inductive coding categories. The master list replaced the *a priori* template for categorisation of the measures during data collection. It was shared with the data collectors via a Google spreadsheet and updated daily.

A wide range of different public sources were used to populate, update and curate our dataset, including official government sources, peer-reviewed and non-peer-reviewed scientific papers, webpages of public health institutions (WHO, CDC, and ECDC), press releases, newspaper articles, and government communication through social media. We collected data on the following: (i) country, (ii) region (when measures were implemented at subnational-level), (iii) date of implementation of the measure, (iv) implemented measures coded following the four-level classification scheme described above (theme, category, subcategory and code), and (v) source. For each country, data were preferentially collected in the language of the country by native data collectors (i.e. Austria, Belgium, Bosnia and Herzegovina, Brazil, Canada, Croatia, Czech Republic, Ecuador, El Salvador, France, Germany, Honduras, Hong Kong, India, Italy, Kazakhstan, Kosovo, Kuwait, Mauritius, Mexico, Montenegro, North Macedonia, New Zealand, Portugal, Ireland, Romania, Serbia, Spain, Syria, Taiwan, and United Kingdom). If this was not possible, Google Translate was used to translate documents^25^.

## Data Records

A static copy of the dataset has been archived in figshare, including all NPIs recorded as of time of submission (4 May 2020), spanning the period 31 December 2019 to 25 April 2020 and covering 53 countries over all continents. A dynamic version of the dataset, which will be continually updated, can be accessed via Github: https://github.com/amel-github/covidl9-interventionmeasures) or from Google Drive: https://drive.google.com/open?id=1041U8iWPDSGI6KHIn9Dg7THkXlo3-gui. in CSV format. Each of the rows represents a single individual NPI and is identified by a unique ID. The master list of coding categories is also available from these repositories. We have also established a Github repository available at: https://github.com/amel-github/CCCSL-Codes and provide codes for importing and visualising the data into R statistical software^26^. Furthermore, for purposes of transparency of data collection and to motivate collaborative validation process as well as a large use and development of our dataset, an open library is available, that contains all sources used to collect the data: https://www.zotero.org/groups/2488884/cccsl covid measure project. In order to leverage on the potential of crowdsourcing for populating and curating the CCCSL dataset, we have launched a webpage dedicated to this project at: http://covidl9-interventions.com/ where contributors can fill up a Google form at: https://bit.lv/2KsY0Tn. if they wish to correct entries, add a measure, and/or provide a feedback.

The dataset contains the following fields:

**ID** –Unique identifier for the implemented measure. ID is also used in the Google Form to report erroneous entries. **Country**-The country where the NPI measure was implemented. **IS03** –Three-letter country code as published by the International Organization for Standardization. **Region** –Subnational geographic area (e.g. region, department, municipality, city) where the NPI measure has been locally implemented (i.e. the measure was not implemented nationwide as of the mentioned date). The country name otherwise (i.e. measure implemented nationwide). **Date** –Date of implementation of the NPI. Date of announcement was used when the date of implementation of the NPI could not be found and this was specified in the field *Comment*. **L1_Measure** –Themes (L1 of the classification scheme). Eight themes were defined (see Table 1). **L2_Measure** –Categories (L2 of the classification scheme). Table 1 provides the list of the categories for each theme. **L3_Measure** –Subcategories (L3 of the classification scheme). Provides detailed information on the corresponding category (L2). **L4_Measure** –Codes (L4 of the classification scheme). Corresponds to the finest level of description of the measures. **Comment** –Provides the description of the measure as found in the text data source, translated into English. This field allows to judge the quality of the label for the different levels of the coding scheme and enables to re-assign the measure to the correct theme/category/subcategory/code in case of error or misinterpretation of the data collector^27^. **Source** –Provides the reference for each entry. Enables to trace back potential changes in the meaning of the label during the translation ^27^. Enables to access the description of the measures in the source language and/or to access to the information as it was dispatched originally.

At the time of submission, the CCCSL dataset included information for 3,274 intervention measures, from 53 countries, including 32 European countries, 12 Asian countries, five South American, one North American country, one Oceanian country, one African country, and the Diamond Princess cruise ship. Fig. 1, Table 2, and Table 3 summarize the dataset. A description of the measures grouped per themes (L1) for each country can be computed from the codes available in the Github (https://github.com/amel-github/CCCSL-Codes).

**Fig. 1.**
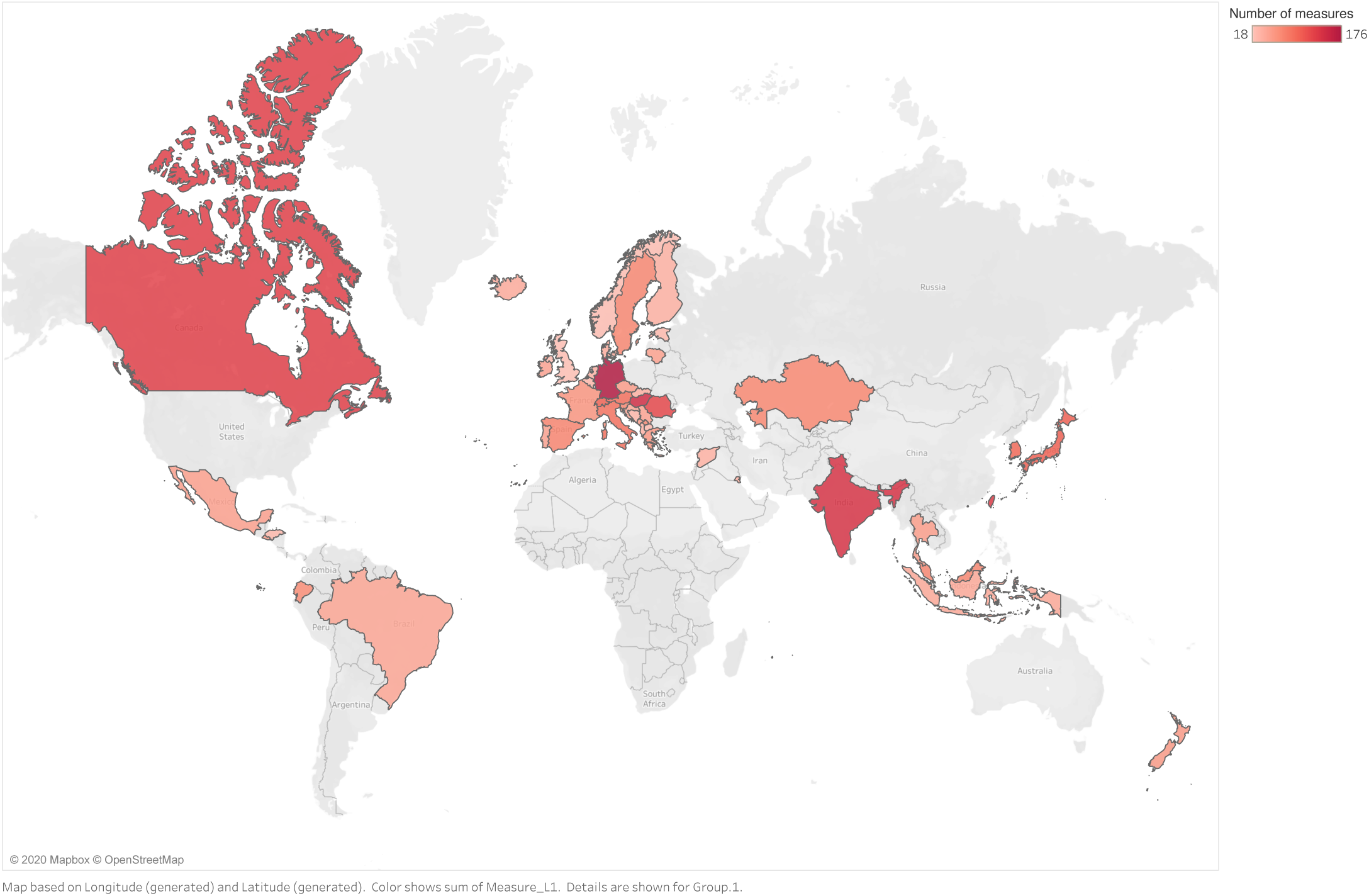
Geographical coverage of the CCCSL and total number of recorded NPIs that were implemented in each country to control the spread of COVID-19. As of date of submission, the dataset includes 53 countries (including the Diamond Princess cruise ship) and dates of NPI implementation range from 31/12/2019 to 25/04/2020.

**Table 2.**
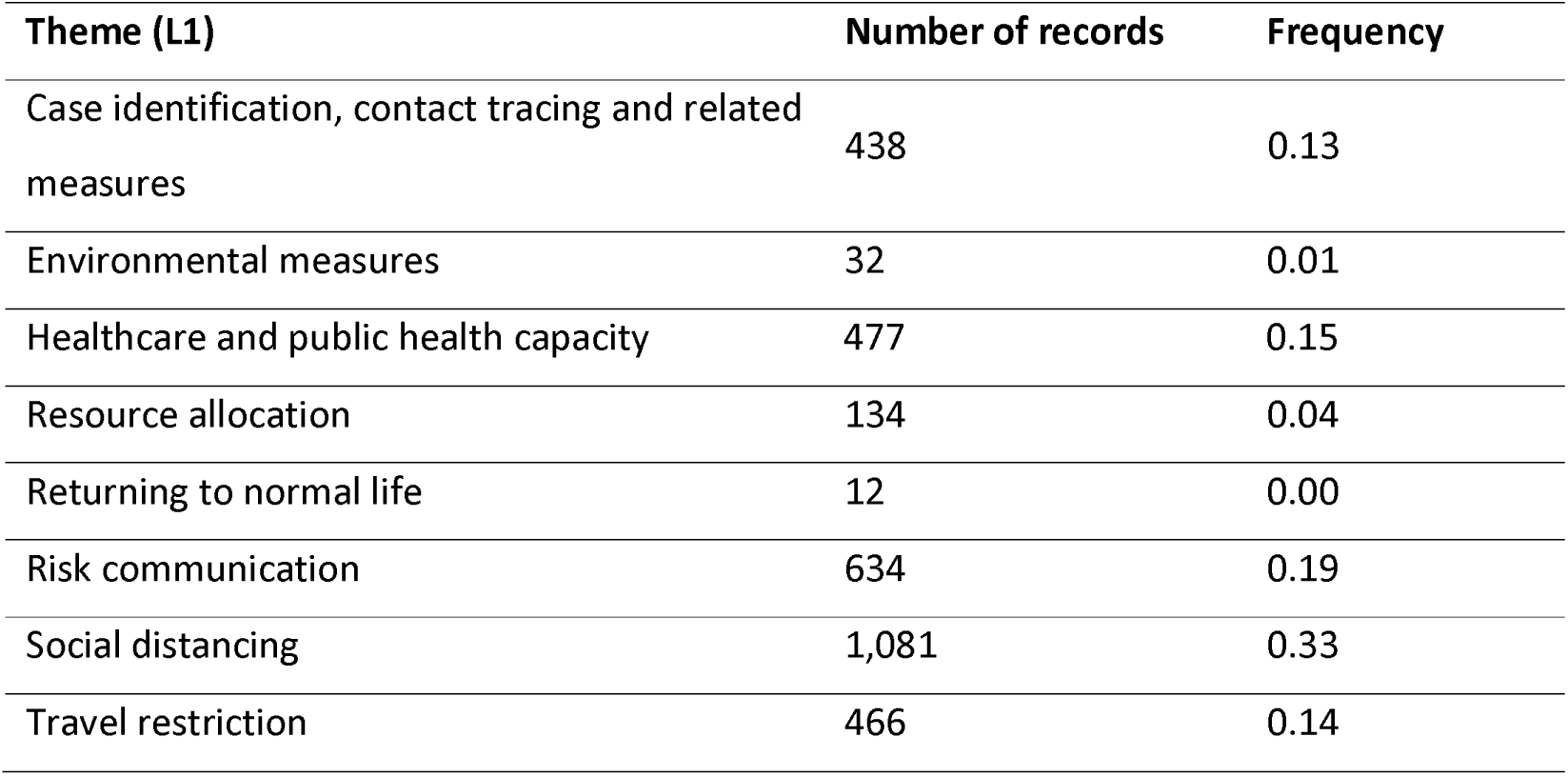
Summary of the government interventions recorded in the CCCSL at level 1 (themes) of the coding scheme. As of date of submission, the dataset includes 53 countries (including the Diamond Princess cruise ship) and dates of NPI implementation range from 31/12/2019 to 25/04/2020.

| Theme (L1) | Number of records | Frequency |
| --- | --- | --- |
| Case identification, contact tracing and related measures | 438 | 0.13 |
| Environmental measures | 32 | 0.01 |
| Healthcare and public health capacity | 477 | 0.15 |
| Resource allocation | 134 | 0.04 |
| Returning to normal life | 12 | 0.00 |
| Risk communication | 634 | 0.19 |
| Social distancing | 1,081 | 0.33 |
| Travel restriction | 466 | 0.14 |

**Table 3.** Top 20 most frequent categories (L2) of NPIs implemented to control the spread of COVID-19. As of date of submission, the dataset includes 53 countries (including the Diamond Princess cruise ship) and dates of NPI implementation range from 31/12/2019 to 25/04/2020.

| Rank | Category (L2) | Number of records | Frequency | Examples of associated subcategories (L3) |
| --- | --- | --- | --- | --- |
| 1 | Educate and actively communicate with the public | 501 | 0.15 | Communication via social media, TV, radio; Campaign for encouraging hand hygiene; Campaign for discouraging gathering; Campaign for respecting respiratory etiquette; Campaign for encouraging self-initiated quarantine; Campaign to promote the 2m distance. |
| 2 | Mass gathering cancellation | 375 | 0.11 | Cancellation of culture events; Cancellation of indoor activities (e.g. fitness gyms, swimming pools, music classes); Closure of place of worship; Sport events; Festivals, faith-based events; Requirement on the number of persons allowed. |
| 3 | Closure of educational institutions | 236 | 0.07 | Kindergartens; Schools; Colleges; Universities; Precise complete or partial closure. |
| 4 | Small gathering cancellation | 205 | 0.06 | Closure of restaurants, bars, cafes; Closure of non-essential shops; Mandatory home office; Family celebrations prohibited; Complete prohibition of gathering; Police and army control and sanctions; Precise the number of persons allowed. |
| 5 | Border restriction | 140 | 0.04 | Borders closed for people from China; Borders closed for people from high-risk areas other than China; Borders closed to all non-citizens; Borders closed for travellers but open for food supply; Borders closed to refugees; Border control. |
| 6 | Quarantine | 125 | 0.04 | Asymptomatic carriers; Contacts; Suspected cases; People who travelled to China or high-risk areas. |
| 7 | Measures for special populations | 121 | 0.04 | Measures to limit contact with the elderly; More exposed professionals (not healthcare), e.g. post officers, public transport drivers; Persons in prison; Vulnerable persons (e.g. homeless). |
| 8 | Crisis management plans | 98 | 0.03 | Develop all-of-society and business continuity plans; Implement repurpose community services plans; Implement repurpose government plans; Research funding. |
| 9 | Enhance emergency response mechanisms | 93 | 0.03 | Set up crisis unit (the mode is described for each government). |
| 10 | Enhance detection system | 83 | 0.03 | Broaden definition / special definition (L4: "of suspected cases"); Research and test of contacts; Research and test of suspected cases; Search for and/or monitor contacts; Test people who travelled in high-risk areas; Test vulnerable population. |
| 11 | Airport health check | 80 | 0.02 | Measure of the temperature; Test of all incoming people with fever or symptoms; Health screening; Health certificate requested. |
| 12 | Individual movement restrictions | 75 | 0.02 | Curfew; Non-essential movements forbidden. |
| 13 | Airport restriction | 67 | 0.02 | Denial entry of travellers from China; Denial entry of travellers from high-risk countries other than China; Specific health channel for travellers from high-risk areas; Denial entry of travellers from Wuhan. |
| 14 | Increase availability of PPE | 65 | 0.02 | Prohibition of export of PPE; Increase availability for healthcare professionals; L3 can precise the type of PPE (face masks, gloves, protective suits) while L4 can precise mean for increasing availability (e.g. import, increase production). |
| 15 | Increase patient capacity | 59 | 0.02 | Number of beds; Build emergency hospitals; Increase ICU capacity; Increase the number of isolation wards; Postponement of non-essential care and non-urgent operations in hospitals. |
| 16 | Return operation of nationals living in regions at risk | 55 | 0.02 | Precise country/region. |
| 17 | Enhance laboratory testing | 51 | 0.02 | Decrease time between test and result; Increase testing capacity (number tests). |
| 18 | Border health check | 46 | 0.01 | Measure of the temperature; Health screening; Health certificate requested; Health declaration; Test at the border. |
| 19 | Increase healthcare workforce | 44 | 0.01 | Mobilization of domestic resources for health; Exception to work law allowed; Train medical staff specially for covid-19; Travelling ban for healthcare professionals; Ease transport for health workers. |
| 20 | Travel alert | 40 | 0.01 | Level 1; Level 2; Level 3 (concerned country is specified) |

## Technical Validation

After the initial data entry, the dataset was checked manually by the data curators. For each measure, concordance between L1, L2, L3, and L4 was checked. Moreover, the unique combinations of L1, L2, L3, and L4 were extracted and controlled for consistency. Typographical and coding errors were also minimized through a manual process. We also initiated a collaborative curation platform relying on internal and external collaborators who exchanged through Slack, Github, Skype, and via emails. This extended effort enabled us to correct typographical and coding errors, to remove line breaks, and to homogenize the dataset for universal use in different programming languages.

Beyond manual validations, we performed a technical validation step to detect possible duplicates. Using the *dplyr* package for the R Statistical Language^28^, we identified any duplicate entries in the vector composed of country, region, date, and the codes from L1 to L4. Those entries were flagged as possible duplicates and reviewed by hand by two curators, ensuring that the dataset does not contain duplicated entries. An R script to reproduce this step is provided at: https://github.com/amel-github/CCCSL-Codes.

While an important effort has been made for standardizing the records, the four-level-α *priori* coding scheme originally proposed showed limitations. First, the existing classification of NPI measures are discordant ^29-31^. We proposed a classification scheme that best fitted our (emergency) needs and the specificity of the COVID-19 pandemic, but this scheme may be subjected to revisions in the future. Secondly, some NPIs have been uniquely implemented (e.g. the deportation of Chinese workers by the Kazakh government) which complicated the coding and categorisation process.

Access to the information from governmental or official source can be compromised if not performed timely. Indeed, several governments or national health agencies update their webpage to provide the latest information to the public. Therefore, access to previous content (i.e. previous restrictions and measures) from official sources was sometimes difficult and some links to online document sources may not exist in the future. Furthermore, while native speakers were recruited whenever possible for data collection, transliteration or translation errors may have occurred when extracting data from Google Translate translations.

We aim to maintain the quality level of the dataset with weekly updates on the countries currently described. Furthermore, we plan to increase the geographic coverage of the dataset, prioritizing large countries (e.g. China, USA, and Australia), those with a high number of reported cases (e.g. Vietnam, Iran, Turkey, Russia, Israel, Peru, Chile Poland, Pakistan, Philippines, Saudi Arabia), and those in which the epidemic is rising and which suffer from a data gap (i.e. African and South American countries). The same technical procedures and the classification scheme described above will be applied to any new information to be included in the dataset. Future versions will be subjected to extensive data validation processes. Four main factors will be considered in future updates: (1) subnational level data; (2) planned duration of measure at time of implementation; (3) new measures versus extension of a previous measure; and (4) measures related to returning to normal life (lifting of restrictions). We plan to stabilize our hierarchical coding scheme for NPIs implemented to contain COVID-19 within six months (measures related to the lifting of the restrictions and adaptive measures that accompany them will be included).

## Usage Notes

The aim of this work is not only to improve the current knowledge on country-based intervention measures implemented to mitigate the burden of COVID-19, but also to characterize the political, public health, and economic strategies of the governments worldwide. Combined with publicly available data on the number of confirmed cases, recovered cases, and deaths ^32^,^33^, the CCCSL dataset makes it possible to assess the effectiveness of the control policies on the COVID-19 epidemic, e.g. the epidemic growth rate or the daily reproduction numbers. The dataset can further benefit the risk assessment of lifting some restrictions and the development of exit strategies. It can also become an essential data source in the aftermath of the first wave of COVID-19, to guide government control policies anticipating a potential second wave of cases. We envision the CCCSL dataset to become a timely valuable and long-lasting data source for assessing the impact of the NPIs on global public health indicators, the economy, and human rights, among others. We provide below three examples of data usage that give an insight into the responsiveness and aggressiveness of the governments in their management of the COVID-19 crisis. In these examples, we excluded data about the Diamond Princess cruise ship, as it represents a special case, and NPIs related to "returning to normal life", as the collect of those measures is ongoing.

### Identify worldwide decision patterns over time

We propose a visualisation of the global decision pattern over 52 countries using a network analysis of the NPIs recorded in the CCCSL at level 2 (categories) of the coding scheme (Fig. 2). The network analysis is based on the overlap in time of the implemented government interventions. Nodes represent NPIs; the size of each node is proportional to the frequency of the NPI (number of countries that implemented this NPI over all considered countries). The network links are directed; the source of a link corresponds to the measure that was implemented first. An interactive version of Fig. 2 is available online at: http://covid19-interventions.com/Network/interventionsNetwork.html. On the panel on the left side of the online Fig. 2, for a selected node (measure), the weight of each link (average overlap in time) is given.

**Fig. 2.**
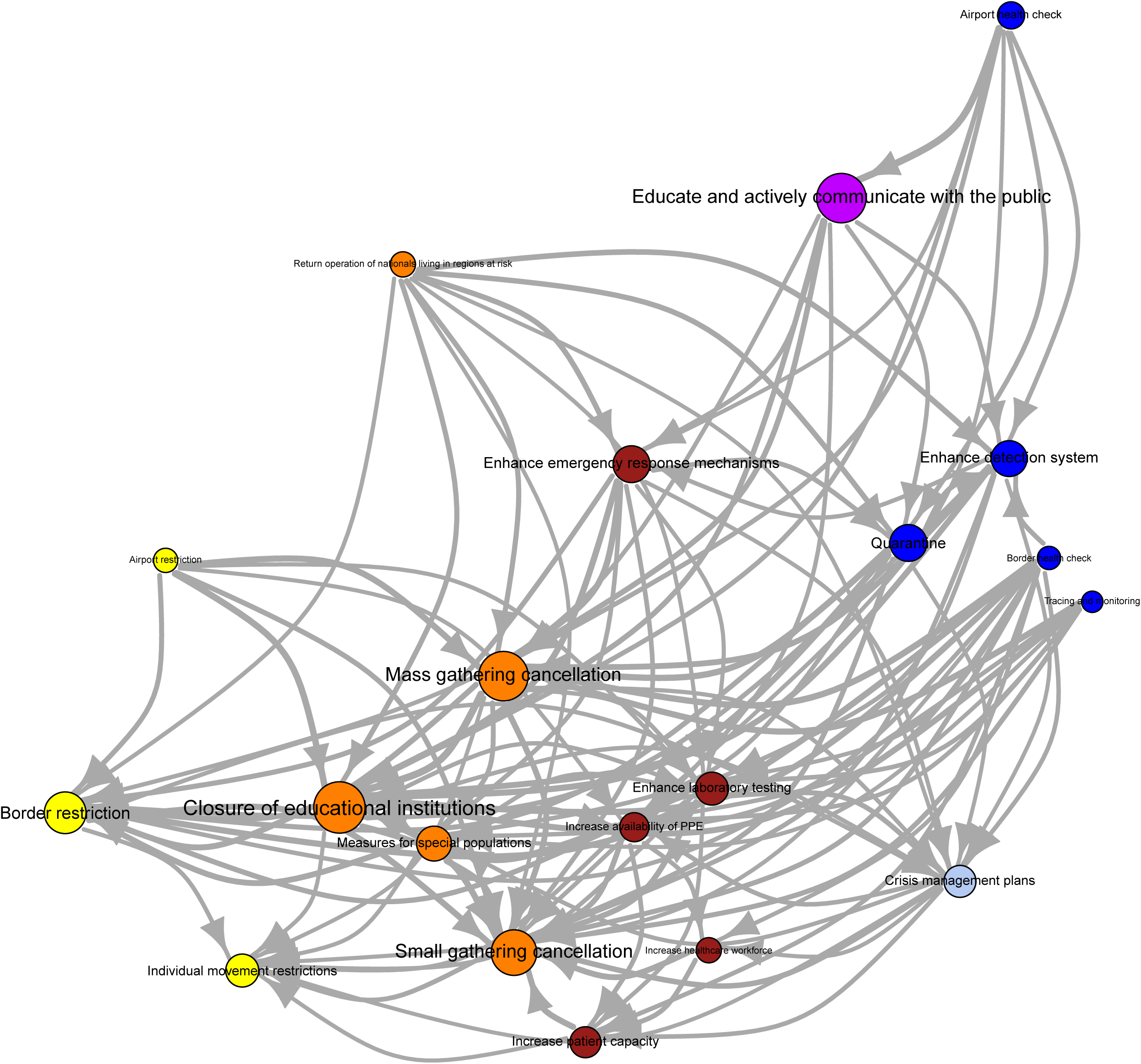
Network of global government decision patterns over time, built using NPIs recorded in the CCCSL at level 2 (categories). The network analysis includes 52 countries (Diamond Princess excluded from this analysis); dates of NPI implementation range from 31/12/2019 to 25/04/2020. The size of the nodes is proportional to the frequency of a given NPI (number of countries that implemented this NPI over all considered countries). The network is filtered such that only edges with weight greater than 0.5 are displayed.

### Mapping the timeline of government interventions through the epidemic

We propose to visualise the time-series of the date of implementation of the NPIs recorded in the CCCSL at level 2 (categories) in the 52 countries (Diamond Princess excluded) using a heat map (Fig. 3). To highlight country-based differences in the timeline of implementation, we used the epidemic age instead of calendar time. For a given day, t, in a certain country, the epidemic age is defined as the time difference, t-t_0_, measured in days, where t_0_ is the first day when the number of confirmed cases was greater or equal to 10. The time-series data of the number of COVID-19 cases was retrieved from the COVID-19 Data Repository by the Center for Systems Science and Engineering (CSSE) at Johns Hopkins University ^33^.

**Fig. 3.**
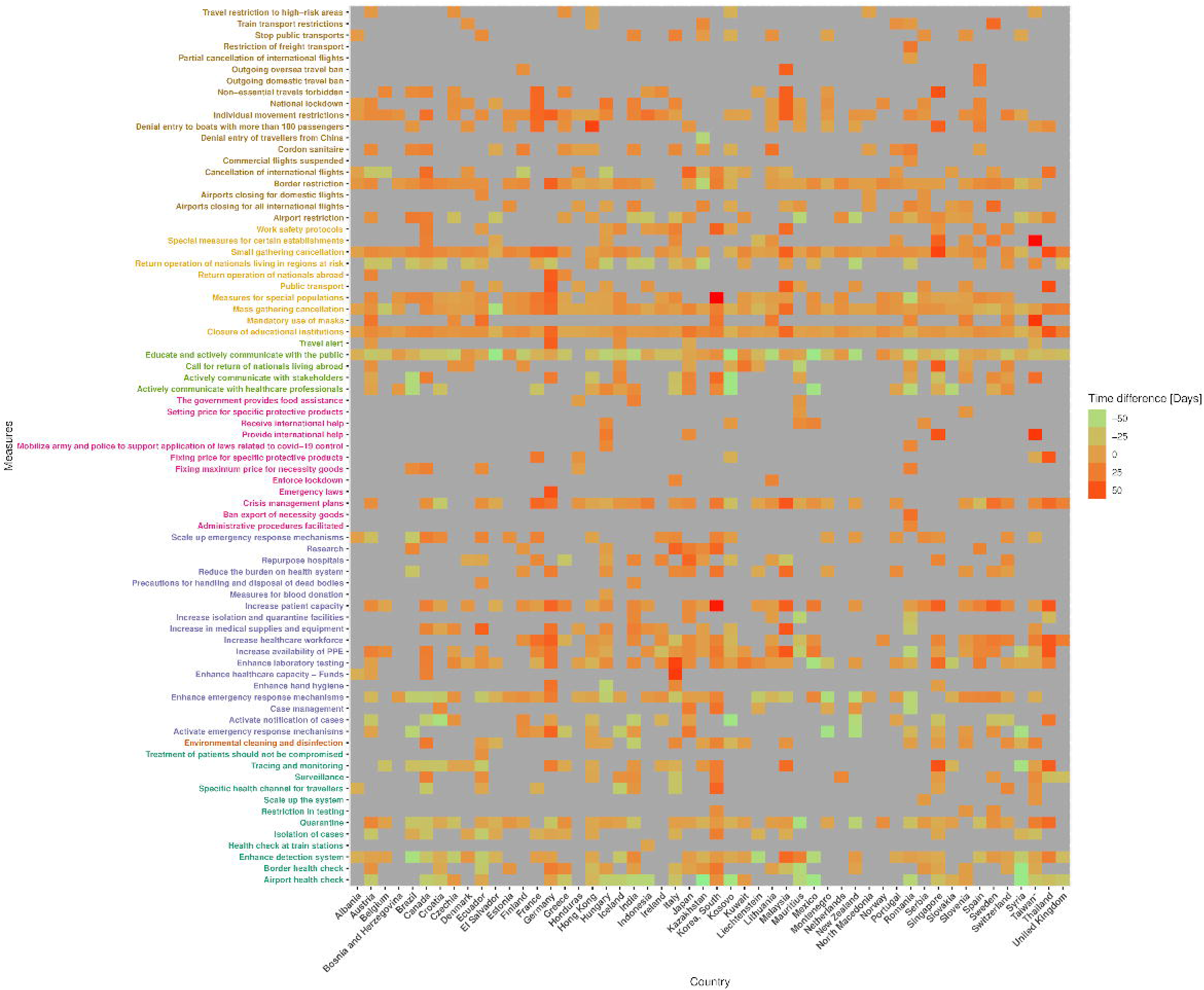
Heat map of the date of implementation of the NPIs recorded in the CCCSL at level 2 (categories) in 52 countries. Time is in epidemic age with t_0_ = day when 10 cases were reported (http://covidl9-interventions.com/CountrylVleasuresHeatmap.svg).

### Country-cluster analysis of the government control strategies

In order to partition the countries based on the aggressiveness (number of NPIs) and responsiveness (timeline) of their control strategy, we applied a k-means clustering. We focused on mandatory government interventions (i.e. the theme "Risk communication" was not included) recorded in the CCCSL at level 2 (categories) that appeared in at least 15 countries, leading to a total number of 29 categories. The clustering algorithm uses the date of implementation of the measures in each country to build a feature vector based on the epidemic age (see above). We considered "anticipatory measures" as those implemented before day when 10 cases were reported; "early measures" as those implemented at the beginning of the epidemic, i.e. between the day when 10 cases were reported and the day when 200 cases were reported; and "late measures" as those implemented at a later stage of the epidemic, i.e. after the day when 200 cases were reported. The algorithm takes also into account the number of measures implemented at these different stages of the epidemic. The time-series data of the number of COVID-19 cases was retrieved from the COVID-19 Data Repository by the Center for Systems Science and Engineering (CSSE) at Johns Hopkins University ^33^. The optimal number of cluster, k, determined by optimizing the within-cluster sums of squares, was seven, explaining 80.3% of the variance (Fig. 4). An interactive version of Fig. 4 is available online at: http://covidl9-interventions.com/CountryClusters.html.

**Fig. 4.**
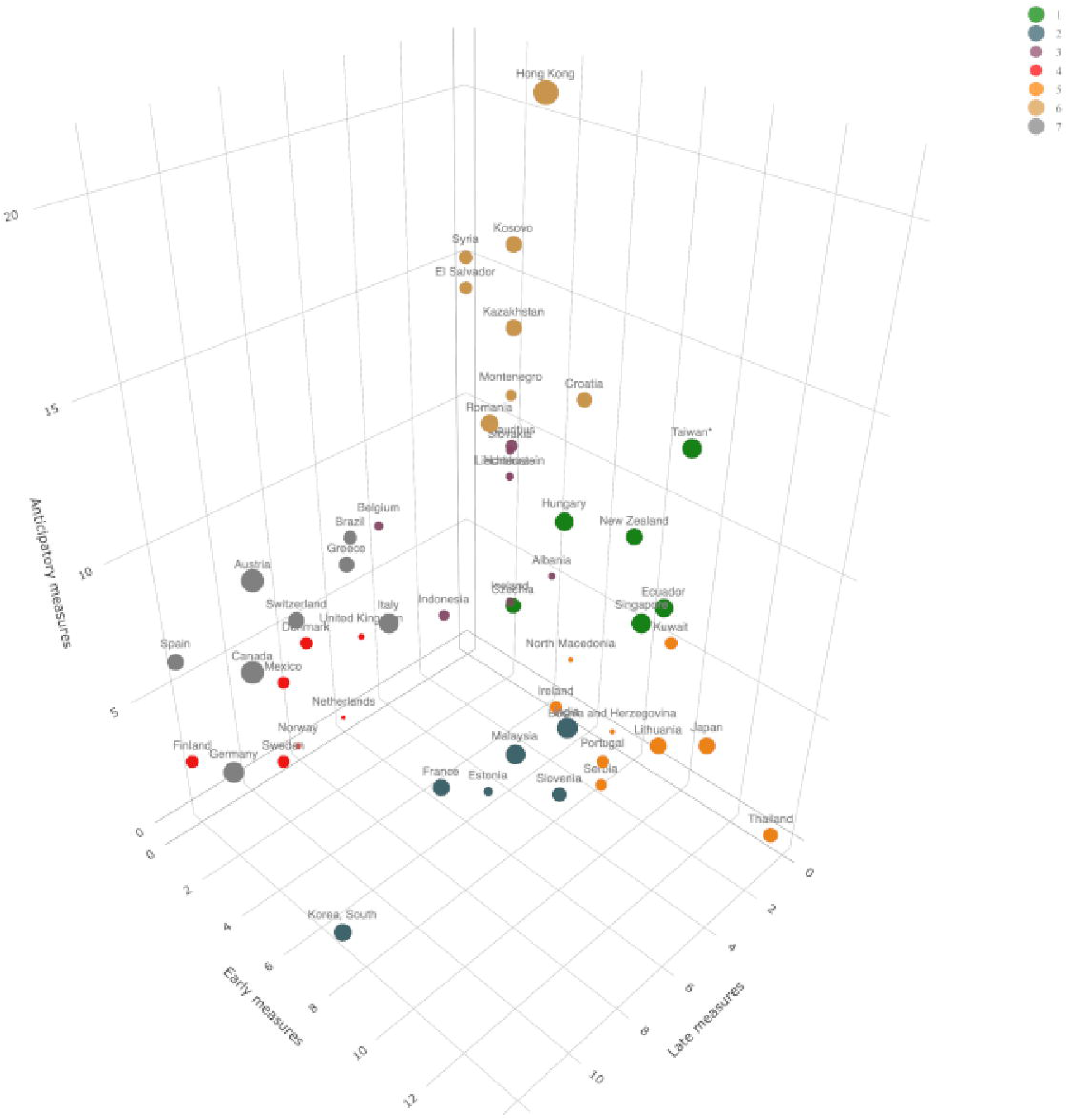
Country-cluster analysis based on the number of mandatory government interventions and respective dates of implementation, as calculated using the epidemic age (t_0_ = day when 10 cases were reported).

## Data Availability

A GitHub record of this project is accessible at https://github.com/amel-github/covid19-interventionmeasures. The codes used to describe the CCCSL dataset and the codes used to explore the CCCSL dataset in the Usage Notes section are available at: https://github.com/amel-github/CCCSL-Codes.

https://github.com/amel-github/CCCSL-Codes

## Code Availability

A GitHub record of this project is accessible at https://github.com/amel-github/covidl9-interventionmeasures. The codes used to describe the CCCSL dataset and the codes used to explore the CCCSL dataset in the Usage Notes section are available at: https://github.com/amel-github/CCCSL-Codes.

## Acknowledgements

D.G. and A.D.N. acknowledge funding from the Vienna Science and Technology Fund - WWTF (VRG16-005). The authors acknowledge William Schueller for his help in the recruitment of the team of data collectors and Petar Sekulic for his advises in the analysis of the dataset. We warmly thank Caspar Matzhold and Michaela Kaleta for checking and testing our code components.

## Author contributions

A.D.L. managed and coordinated the production of the dataset and prepared it for publication, including: developing the coding scheme, collecting and curating the data, managing the team of data collectors, creating the library of sources, writing the data descriptor, and creating the tables. The following authors: E.D., N.H., T.N., E.C., A.D.N., J.L., D.S.G., and A.R. produced a substantial work to generate the dataset and prepare it for publication, including: developing the coding scheme, collecting the data, formatting the data for presentation of the published work, visualising the data, and creating the library of sources. D.G. created the webpage dedicated to this project. The following list of authors (listed alphabetically): A.C., A.T., A.D., A.P., D.C., D.L., D.B., E.F.T., H.T., J.A., J.K., J.R., J.S., L.H., L.S., L.G., L.G., M.R.F., R.V., S.H., S.Á., S.L., S.H., V.A., V.R., V.D.P.S., X.C. and X.M.P.O. includes the many individuals who collected and curated data through the dedicated platform and provided comments to facilitate the use of the dataset across different programming platforms. A.D.L. and D.G. jointly supervised this work. S.T. mentored the core team. All authors reviewed and approved the final manuscript.

